# A Generalizable Data Assembly Algorithm for Infectious Disease Outbreaks

**DOI:** 10.1101/2021.04.21.21255862

**Authors:** Maimuna S. Majumder, Sherri Rose

## Abstract

**Background & Objective:** During infectious disease outbreaks, health agencies often share text-based information about cases and deaths. This information is usually text-based and rarely machine-readable, thus creating challenges for outbreak researchers. Here, we introduce a generalizable data assembly algorithm that automatically curates text-based, outbreak-related information and demonstrate its performance across three outbreaks.

**Methods:** After developing an algorithm with regular expressions, we automatically curated data from health agencies via three information sources: formal reports, email newsletters, and Twitter. A validation data set was also curated manually for each outbreak.

**Findings:** When compared against the validation data sets, the overall cumulative missingness and misidentification of the algorithmically curated data were ≤2% and ≤1%, respectively, for all three outbreaks.

**Conclusions:** Within the context of outbreak research, our work successfully addresses the need for generalizable tools that can transform text-based information into machine-readable data across varied information sources and infectious diseases.

## Introduction

Since 2000, thousands of infectious disease outbreaks have been reported by the World Health Organization (WHO) globally [1]. A considerable subset of these have been due to emerging zoonotic pathogens, including the novel coronavirus SARS-CoV-2, the causative agent of the Coronavirus Disease 2019 (COVID-19); its predecessors, Middle East Respiratory Syndrome (MERS) coronavirus and SARS-CoV-1; Zika virus, and Ebola virus, among others [2–4]. Emergence of these pathogens has been driven by the increasing permeability of the animal-human interface, whereas ease of travel has enabled their transmission across borders [5,6]. Not all outbreaks from the last two decades have been due to emerging infections, however; notably, due to increasing vaccine hesitancy around the world, *re*-emerging diseases, such as measles and mumps, have experienced a resurgence as well [7,8].

During these outbreaks, epidemiological information from a variety of data sources—from formal reports by the WHO to email newsletters and social media posts from national ministries of health—is often made available to the public, including researchers responsible for monitoring and mitigation efforts [1,9–14]. Unfortunately, these publicly available data are typically locked in blocks of text that are rarely machine-readable [15], which poses a considerable roadblock for surveillance and response activities that hinge on mathematical modeling (e.g., data-driven allocation of ventilators or vaccines). To overcome this hurdle, researchers typically commit substantial labor towards manually curating and converting these text-based data into an analyzable format (e.g., comma-separated values, CSV). The time and effort required is often directly related to the complexity of the available information.

In this paper, we introduce a generalizable data assembly algorithm to automate curation of text-based, outbreak-related information and demonstrate its performance across three recent case study outbreaks: measles in Samoa (2019), Ebola in the Democratic Republic of the Congo (DRC) (2018–2019), and MERS in South Korea (2015). We implement this algorithm on source text of increasing complexity from social media (i.e., Twitter), email newsletters, and WHO disease outbreak news (DON) reports, respectively, to produce machine-readable CSV files for each of our three case studies. Though the data available for curation vary across source texts, the underlying structure of the algorithm—regular expressions to extract pertinent outbreak-related information— remains constant across applications and is generalizable.

The source texts considered in this study represent a spectrum of information complexity, and when combined with mathematical modeling approaches, can be used to inform decision-making during infectious disease outbreaks. For measles in Samoa and Ebola in the DRC, we extract simple aggregate statistics (e.g., case counts) over time, which can be used for case count projections, assessment of intervention performance, and vaccination rate estimation [17–30]. Meanwhile, for MERS in South Korea, we extract more complex multi-feature patient-level data (i.e., data in which every row is a patient, and every column is a feature), which enable reconstruction of transmission networks and evaluation of risk factors associated with mortality [31–39].

## Methods

Data on the evolving epidemiology of each outbreak were first manually curated for validation purposes. Summary information for each study is available in Table 1. Aggregate cases and deaths associated with the measles outbreak in Samoa were collected from the Government of Samoa Twitter account from November 22, 2019 (date of first tweet) to December 8, 2019 (date of last tweet) [9,10]. Similar aggregate statistics were also collected for the Ebola outbreak in the DRC from email newsletters issued by the Ministère de la Santé RDC (MSRDC) from August 6, 2018 (date of first newsletter received) to July 31, 2019 (date of last newsletter received) [11,12]. Finally, patient-level data were collected from WHO DON reports for the MERS outbreak in South Korea from May 30, 2015 (date of first report) to June 9, 2015 (date of last report) [13,14]. These same text-based data were then algorithmically collected using our data assembly algorithm.

**Table 1.** Data collected across case study outbreaks.

| **Case Study** | **Data Source** | **Reporting Period** | **Number of Fields** | **Total Cells Curated** |
| --- | --- | --- | --- | --- |
| Measles in Samoa | Twitter | November 22, 2019 – December 8, 2019 | 3 | 51 |
| Ebola in the DRC | Email Newsletters | August 6, 2018 – July 31, 2019 | 10 | 3600 |
| MERS in South Korea | Disease Outbreak News Reports | May 30, 2015 – June 9, 2015 | 5 | 315 |

The assembly algorithm was developed in the Python programming language and, as shown in Figure 1, uses regular expressions and trigger phrases to automatically transform semi-structured text-based information into machine-readable data. Here, trigger phrases are the phrases that accompany the information of interest in a given block of text. When these phrases are translated into searchable patterns of characters (i.e., regular expressions), they act as “triggers” for the data assembly algorithm to identify and collect information for the desired fields (i.e., variables). This underlying regex-based structure enables generalizability of the algorithm to a wide variety of source texts and information types, as demonstrated by the three case study outbreaks selected.

**Figure 1.**
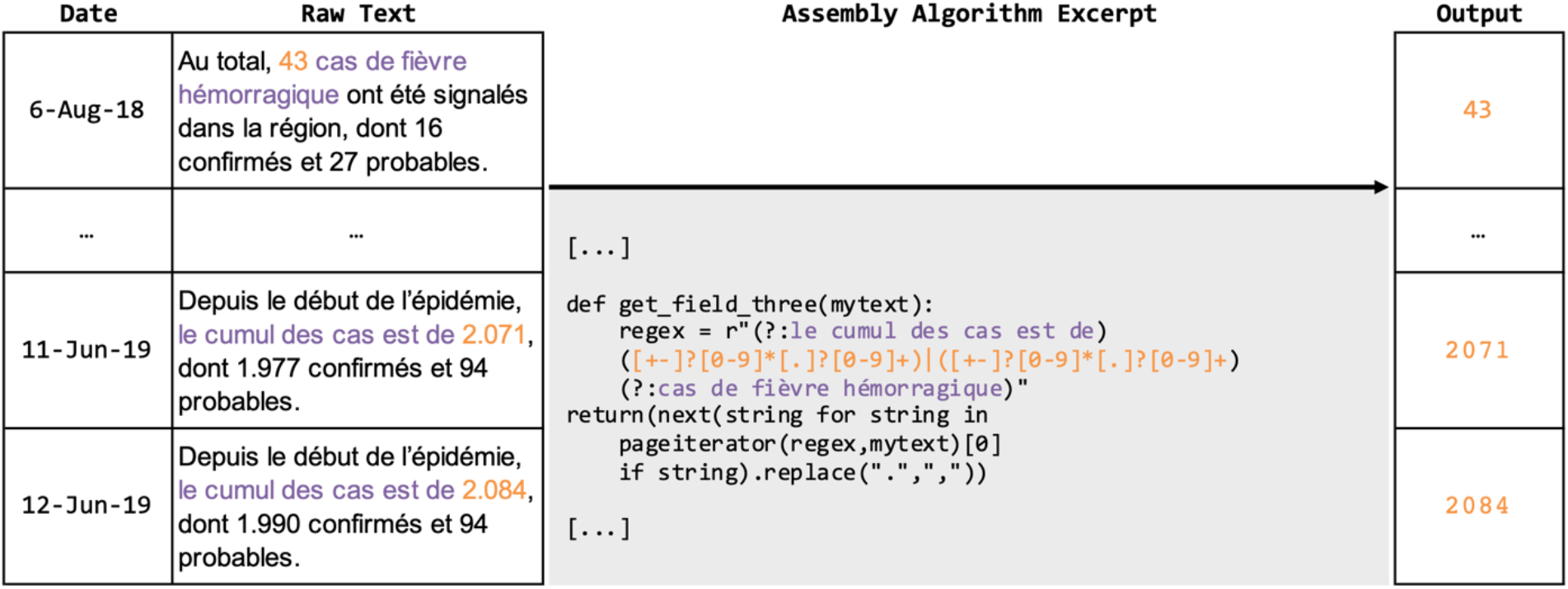
Assembly algorithm flowchart depicting automatic curation of text-based information into machine-readable data. Three example rows of data from the Ebola case study are shown for a single field (of 360 rows and 10 fields total). Trigger phrases are shown in purple and the numerical values of interest are shown in orange.

For the measles case study, the following three data fields were automatically curated using our assembly algorithm: cumulative cases, incident cases, and cumulative deaths. Seventeen rows of data, where each row is a date, were collected across these three fields for a total of 51 cells. Similarly, data for the following 10 fields were automatically curated for the Ebola case study: confirmed cumulative cases, total cumulative cases (confirmed + probable), confirmed cumulative deaths, total cumulative deaths (confirmed + probable), cumulative cases recovered, cumulative vaccinations deployed, cumulative vaccinations deployed in Region A, cumulative vaccinations deployed in Region B, cumulative vaccinations deployed in Region C, and cumulative vaccinations deployed in Region D. Across these 10 fields, 360 rows of data, where again each row is a date, were collected for a total of 3600 cells. Finally, data for the MERS case study were automatically curated to populate the following five fields: documented sex, age, date of symptoms, date of diagnosis, and healthcare worker status. Sixty-three rows of data, where each row is a patient, were collected for a total of 315 cells across these five fields.

In all three case study outbreaks, the manually curated data for the aforementioned fields were used to validate the performance (i.e., missingness and misidentification) of the assembly algorithm. Missingness is defined as a cell for which the algorithm did not curate a value but for which a value was available when compared against manual curation. Misidentification is defined as a cell for which the algorithm curated a value but for which the value was incorrect when compared against manual curation. Given its intended application in outbreak settings, the assembly algorithm was designed conservatively, placing priority on increasing accuracy over decreasing missingness. Code for all three implementations of the assembly algorithm, as well as the manually collected validation data, are available at <https://github.com/mmajumder/Data_Assembly_Algorithm>.

## Results

When validating algorithmically collected data against manually collected data, the data assembly algorithm performed well for all three iterations. Across the entirety of each outbreak reporting period, overall cumulative missingness for the case studies was 0% (0 cells) for measles, 1% (34 cells) for Ebola, and 2% (7 cells) for MERS, while overall cumulative misidentification was 0% (0 cells), 0% (0 cells), and 1% (3 cells), respectively.

Because the reporting period for the Ebola outbreak was considerably longer (368 days) than the measles (16 days) and MERS (11 days) case studies, we also examined missingness and misidentification over time by day for the Ebola case study. Notably, the assembly algorithm exhibited steady gains in cumulative accuracy from August 2018 through June 2019, as displayed in Figure 2. Decreased cumulative availability of data in the source itself (i.e., fields for which MSRDC reported data in May 2019 but no longer reported in June 2019) coincided with minor decreases in cumulative accuracy between June 2019 and August 2019. Cumulative missingness dropped from 5% in August 2018 to near 0% in August 2019, and due to the conservative nature of the assembly algorithm, cumulative misidentification was 0% over the same time period.

**Figure 2.**
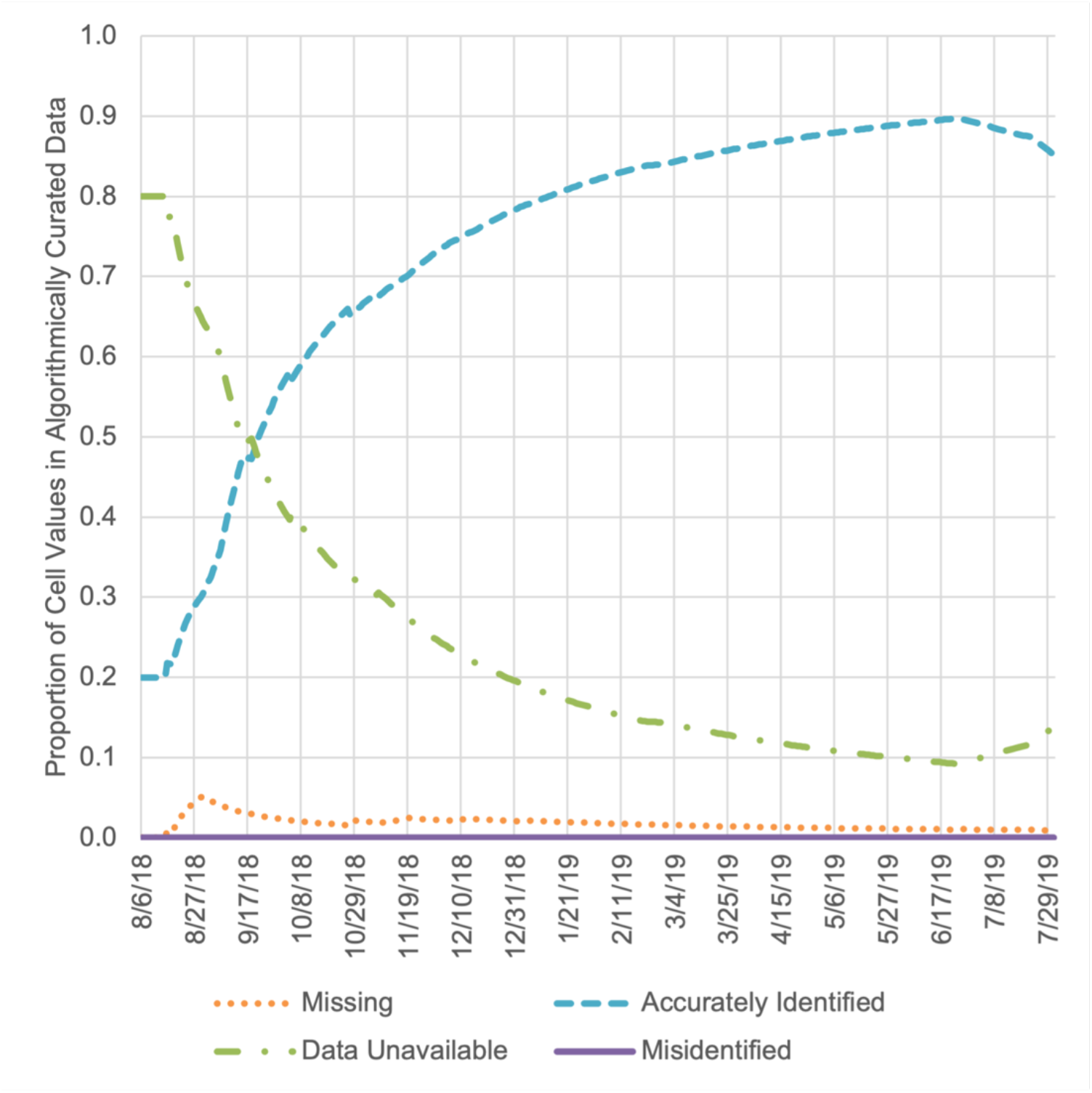
Assembly algorithm performance curves over time for the Ebola case study. Cumulative missingness is shown in orange, accuracy in teal, misidentification in purple, and data availability (at the source) in green.

## Discussion

By showcasing its performance within the context of three distinct infectious disease outbreaks, we demonstrated the generalizability of our data assembly algorithm across diverse source texts and information types. Intuitively, we found that algorithmic curation of more complex data (e.g., multi-feature patent-level data for MERS in South Korea) exhibited slightly higher rates of missingness and misidentification than simpler data (e.g., case counts over time); however, overall cumulative performance for both metrics was impressive across curated fields for all three case study outbreaks.

The fields for which data were automatically curated by our assembly algorithm were selected purposefully given their long-standing utility to mathematical modeling for informed epidemiological decision-making. Historically, counts of cases and deaths over time—fields that were collected both for measles in Samoa and for Ebola in the DRC—have been used to model the transmission dynamics associated with outbreaks, including important epidemiological parameters such as fatality rates and reproduction numbers [17–30,40–45]. These parameters are critical to formulating case count projections [17– 21] and assessing performance of interventions [22–25], which enable public health decision-makers to approach outbreaks from a position of preparedness. Furthermore, these parameters can also be used to model vaccination rates during outbreaks of vaccine-preventable diseases, which can be leveraged to lobby for the resources necessary to vaccinate vulnerable communities [26–30]. Meanwhile, patient-level “line list” data have traditionally been employed to assess risk factors for different outcomes [31–38]; indeed, the data presented in this paper for MERS in South Korea have been used precisely in this way to assess risk factors for mortality given MERS-CoV infection [31,32], as well as for transmission to others following infection [38]. Such analyses allow for improvements to resource allocation both with respect to patient care (i.e., preferentially allocate intensive care units to patients who are less likely to survive infection) and with respect to contact-tracing (i.e., preferentially allocate resources to contact trace individuals who are more likely to transmit to others following infection), among other applications.

As recently noted by George et al. [15], tools that can transform text-based information into machine-readable data are urgently needed by the outbreak management community. Given the epidemiological utility of the data types curated by our data assembly algorithm across our three case study outbreaks, we believe that the usefulness of the work we present here will persist as infectious diseases continue to emerge and re-emerge. We encourage other researchers to apply it to novel contexts (i.e., new outbreaks), while carefully considering the ethical implications before deployment in new settings [46]. Our algorithm is designed to generalize across diseases and enable the democratization of essential epidemiological data that are otherwise locked in blocks of non-machine-readable text. However, despite strong accuracy and missingness assessments for all three case study outbreaks considered in this paper, we recommend that random manual checks be implemented to validate the robustness of our data assembly algorithm when employed during future outbreaks.

## Data Availability

Please refer to the manuscript for a link to the study's Github repository.

## Author Contributions

<u>Maimuna S. Majumder, PhD:</u> Study design; data acquisition, analysis, and interpretation; drafting the work; critical revision of the work

<u>Sherri Rose, PhD:</u> Study design; data interpretation; critical revision of the work

